## Supplementary Material for "From Abstainers to Dependent Drinkers: Alcohol Consumption Patterns and Risk Factors Among Portuguese University Students"

### Contents

Supplementary Figures ........................................................................................ 2

Results for Group Differences of the ACQ-SF-R Factor Scores ................................ 4

### Supplementary Figures

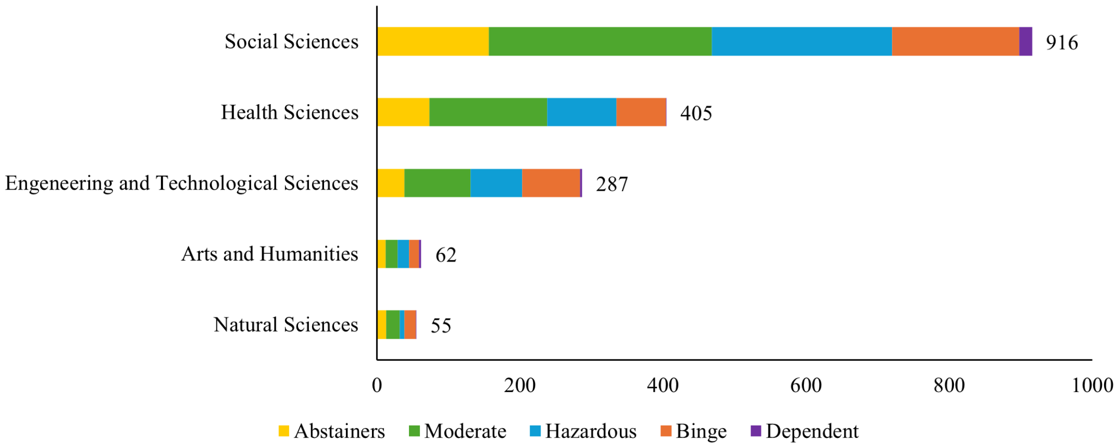


**Figure S1.** Distribution of Drinking Groups by Study Area.

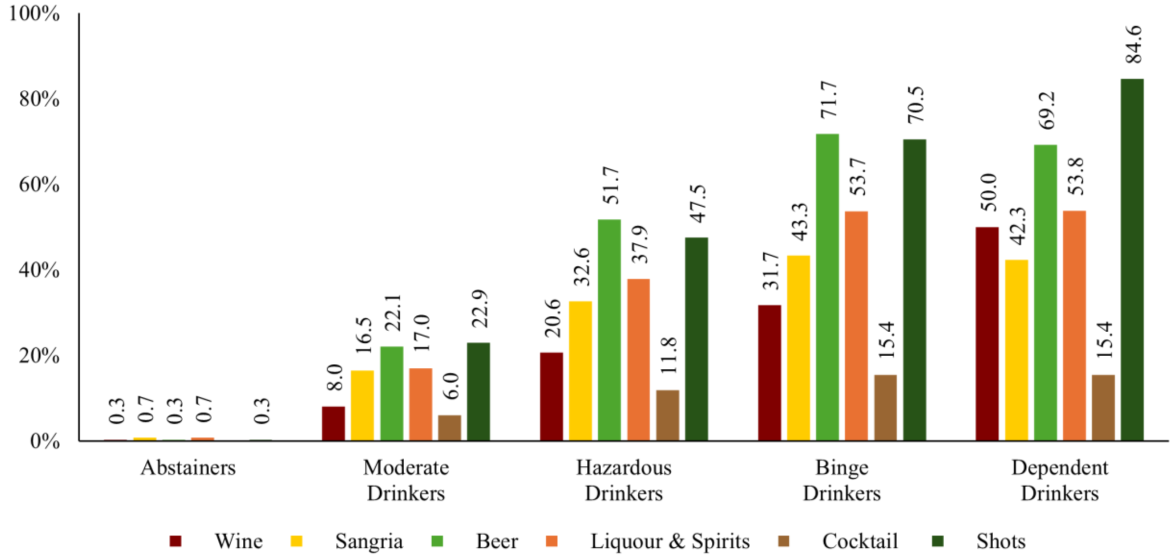


**Figure S2.** Distribution of type of alcohol consumption by Drinking Groups.

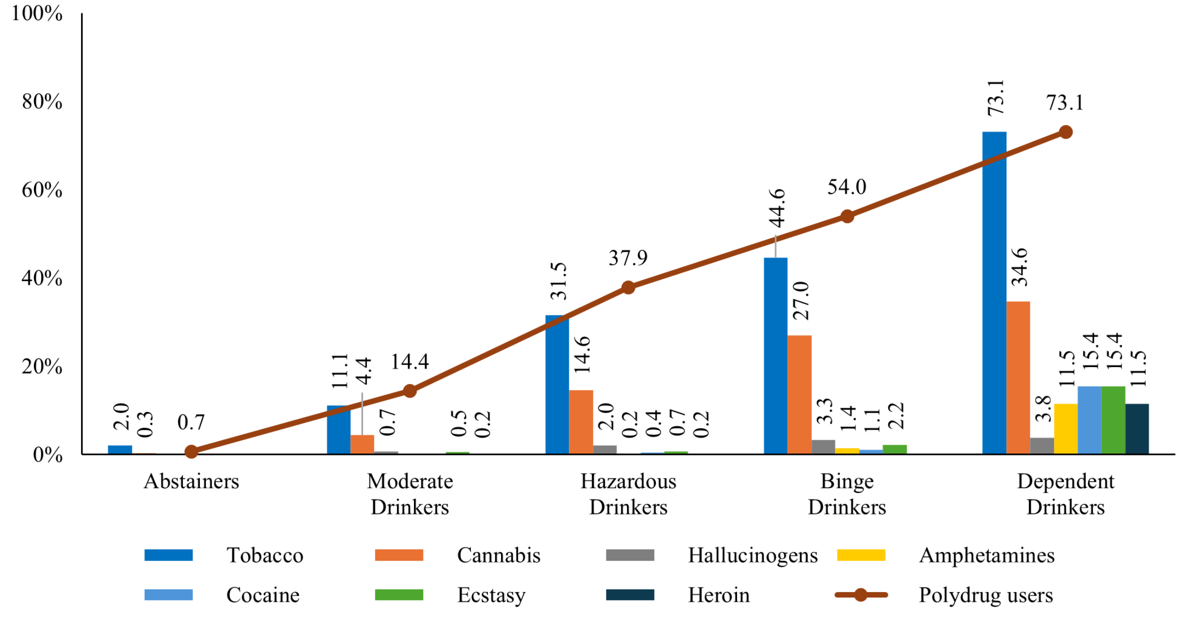


**Figure S3.** Distribution of type of drug use (bar plot), and prevalence of polydrug use (line), across the Drinking Groups.

### Results for Group Differences of the ACQ-SF-R Factor Scores

Regarding the ACQ-SF-R factor scores, significant differences between groups were also revealed for Emotionality [$F(4, 95.821) = 38.837$, $p < .001$, $\omega^2 = .141$], Purposefulness [$F(4, 95.829) = 66.319$, $p < .001$, $\omega^2 = .258$] and Compulsivity [$F(4, 94.954) = 67.564$, $p < .001$, $\omega^2 = .231$].

*Post-hoc* comparisons for the Emotionality factor showed higher scores for Dependent Drinkers, with significant differences compared to Moderate Drinkers ($p = .018$) and Abstainers ($p = .003$). Similarly, Binge Drinkers had significantly higher scores compared to Moderate Drinkers ($p < .001$) and Abstainers ($p < .001$). Hazardous Drinkers also scored higher than Moderate Drinkers ($p < .001$) and Abstainers ($p < .001$), and Moderate Drinkers had higher scores than Abstainers ($p < .001$).

Regarding the Purposefulness factor, the highest scores were found in Dependent Drinkers, with significant differences compared to Moderate Drinkers ($p = .016$) and Abstainers ($p = .001$). Binge Drinkers had higher scores compared to Hazardous Drinkers ($p < .001$), Moderate Drinkers ($p < .001$) and Abstainers ($p < .001$). Hazardous Drinkers scored higher than Moderate Drinkers ($p < .001$) and Abstainers ($p < .001$); and Moderate Drinkers had higher scores than Abstainers ($p < .001$).

Lastly, for the Compulsivity factor, the Dependent Drinkers group displayed the highest scores, with significant differences compared to Hazardous Drinkers ($p = .048$), Moderate Drinkers ($p = .008$), and Abstainers ($p = .003$). Binge Drinkers showed higher scores than Hazardous Drinkers ($p = .016$), Moderate Drinkers ($p < .001$), and Abstainers ($p < .001$). Hazardous Drinkers scored higher than Moderate Drinkers ($p < .001$) and Abstainers ($p < .001$); and Moderate Drinkers had higher scores than Abstainers ($p < .001$).
